# Developing Machine Learning Models for Predicting Intensive Care Unit Resource Use During the COVID-19 Pandemic

**DOI:** 10.1101/2021.03.19.21253947

**Authors:** Stephan Sloth Lorenzen, Mads Nielsen, Espen Jimenez-Solem, Tonny Studsgaard Petersen, Anders Perner, Hans-Christian Thorsen-Meyer, Christian Igel, Martin Sillesen

## Abstract

**Importance:** The COVID-19 pandemic has put massive strains on hospitals, and tools to guide hospital planners in resource allocation during the ebbs and flows of the pandemic are urgently needed.

**Objective:** We investigate whether Machine Learning (ML) can be used for predictions of intensive care requirements 5 and 10 days into the future.

**Design:** Retrospective design where health Records from 34,012 SARS-CoV-2 positive patients was extracted. Random Forest (RF) models were trained to predict risk of ICU admission and use of mechanical ventilation after *n* days (*n* = 5, 10).

**Setting:** Two Danish regions, encompassing approx. 2.5 million citizens.

**Participants:** All patients from the bi-regional area with a registered positive SARS-CoV-2 test from March 2020 to January 2021.

**Main outcomes:** Prediction of future 5- and 10-day requirements of ICU admission and ventilator use. Mortality was also predicted.

**Results:** Models predicted 5-day risk of ICU admission with an area under the receiver operator characteristic curve (ROC-AUC) of 0.986 and 5-day risk of use of ventilation with an ROC-AUC of 0.995. The corresponding 5-day forecasting models predicted the needed ICU capacity with a coefficient of determination (R^2^) of 0.930 and use of ventilation with an R^2^ of 0.934. Performance was comparable but slightly reduced for 10-day forecasting models.

**Conclusions:** Random Forest-based modelling can be used for accurate 5- and 10-day forecasting predictions of ICU resource requirements.

**Funding:** The study was funded by grants from the Novo Nordisk Foundation to MS (#NNF20SA0062879 and #NNF19OC0055183) and MN (#NNF20SA0062879).

The foundation took no part in project design, data handling and manuscript preparation.

**KEY POINTS:** *Question:* Can machine learning models (ML) be used for predicting hospital and intensive care unit (ICU) resource requirements, and thus assist in logistics crisis management during the COVID-19 pandemic?

*Findings:* Retrospective study of the resource use of 34.012 COVID-19 patients during the first and second COVID-19 wave in Denmark. ML models were trained for the purpose of predicting the number of patients needing ICU admission and ventilators 5 and 10 day after their first positive SARS-CoV-2 test. The study demonstrates that ML models can accurately predict intensive care admission requirements with 5-day area under the receiver operator characteristic curve (ROC-AUC) of 0.986 and need for ventilator support with a ROC-AUC of 0.995. 10-day predictions were comparable.

*Meaning:* The study demonstrates that ML modelled could be a useful tool for hospital planners during crisis management, including the current COVID-19 pandemic.

## INTRODUCTION

Since the outbreak of the COVID-19 pandemic in early 2020, almost 90 million confirmed cases and 1.9 million deaths have occurred as a result of the SARS-CoV-2 infection worldwide^1^. The speed of viral spread combined with hospital and governmental systems being ill-prepared for large-scale pandemic responses, created a situation where allocation of health care resources, including the mobilization of ad-hoc intensive care and isolation units were urgently needed.

As hospital resources were redirected towards the COVID-19 response, elective visits and planned surgical procedures quickly became the victim of collateral damage induced by the shift in health care resources, resulting in a massive backlog of elective procedures that will likely take months or even years to overcome.

As expected, the pandemic hit societies in waves, creating an ebb during the summertime where resources could again be partly redirected towards other tasks. The choice of when to up or downscale the COVID-19 response thus quickly became a challenge for hospital planners, mobilizing resources at the time of pandemic acceleration while re-routing physical as well as staff-resources during the months characterized by low infection rates.

Deciding when to increase the resources for the pandemic response at the cost of the elective workload is, however, a challenge. Such decisions are dependent on accurate prediction models capable of risk-stratifying patients based on available health information on confirmation of SARS-CoV-2 infection, as well as ensuring that these predictive models can adjust to changes in disease patterns as the pandemic progresses from the first to subsequent waves of infection^2, 3^. And indeed, according to a recent review by Becker et al as of Sept 27, 2020, there have been more than 5000 modelling analyses published in peer-reviewed journals, excluding preprint servers, since the start of the epidemic ^4^. In contrast to mechanistic models, we and others have trained machine learning (ML) models on electronic health record (EHR) data towards risk prediction on SARS-CoV-2 infection, specifically targeting resource-requiring events such as hospitalization, ICU admission and use of mechanical ventilation^5–7^. We found that accurate risk prediction of the individual patient (micro-prediction) can be performed with a limited EHR dataset, thus opening the potential for predicting hospital resource requirements on a population-wide scale (macro-prediction) in advance, based on available EHR data at the time of COVID-19 diagnosis.

Usual epidemiological modelling takes into account the number of infected patients, and, based on a small number of constants, partial differential equations model the future development of the epidemic ^8^. While providing insights into the role of macroscopic variables on the disease dynamics, such approaches have limited ability to model shifts in epidemic constants arising from the underlying demography of the patients as well as changes in the testing strategy. An approach based on the full medical history of all individuals tested positive for SARS-CoV-2 has the potential to overcome these issues as shifts in the epidemic constants will be reflected in the inputs, and by considering the more detailed information provided at the level of individuals – in contrast to macroscopic variables at population level – it may allow for higher accuracy. We hypothesize that such an approach modelling the epidemic severity development based on relevant clinical information can yield more precise forecasting essential for the resource management in hospitals.

Thus, this study investigates whether the previously trained ML models^7^ can be retrained for the purpose of *n*-day forecasting, capable of predicting the number COVID-19 related ICU admissions and use of mechanical ventilation in a bi-regional cohort in Denmark. While the approach is also applicable to hospital admission in theory, patients are often not known to the system in the *n-*day timeframe prior to admission (due to not yet having been tested for SARS-CoV-2), and thus the method will likely be less precise. Similarly, the approach is also applicable to prediction of mortality within the *n*-day period, although forecasting the number of deaths explicitly is less relevant in terms of determining the need for hospital resources. However, for completeness, we investigated the approach for forecasting both hospital admission and mortality and included results in the supplementary material.

While our suggested approach allows for models for any *n*-day timeframe, we chose a 5- and a 10-day timeframe for our evaluation in order to meet the tradeoff between a relevant window for hospital planners with perceived retention of predictive accuracy.

## DATA

Data access and handling was carried out in accordance with national guidelines and regulations. The study was approved by the relevant legal boards: the Danish Patient Safety Authority (Styrelsen for Patientsikkerhed, approval #31-1521-257) and the Danish Data Protection Agency (Datatilsynet, approval #P-2020-320). Under Danish law, these agencies provide the required legal approval for the handling of sensitive patient data, including EHR data, without patient consent.

### Cohort information

We conducted the study on health data extracted from the Danish Capital and Zealand region EHR system, covering approx. 2.5 million citizens in eastern Denmark. The extracted cohort consisted of 34,012 patients tested positive for SARS-CoV-2 in both regions from March 3^rd^, 2020 to January 5th, 2021. Extracted patient data included age, Body Mass Index (BMI), sex, smoker/non-smoker status and diagnoses. Extracted temporal data included time and date of SARS-Cov-2 PCR tests, hospital and ICU admissions (due to COVID-19), use of mechanical ventilation, medications administered, laboratory tests performed, vital signs recorded, and death where applicable.

### Data set construction

For each patient, a timeline was constructed from the time of the first positive SARS-CoV-2 test and until censoring. Patients were censored at the time of death, a negative SARS-CoV-2 PCR test, 30 days without hospitalization (but 30 days after the positive test at the earliest), or at the time of cohort extraction.

From each patient timeline, a set of *snapshots* was constructed, one for every 24 hours. Each patient snapshot consisted of the set of features extracted for the patient: age, sex, BMI, comorbidities (based on the ICD-10 and ATC codes, for a full definition, please see^7^, smoker/non-smoker status, lab tests, and temporal features: time since positive test result, time spent in hospital, time spent in ICU, and time on a ventilator. See Supplementary Table 1 for a complete overview of features included. For each lab test and vital sign feature, we added the following aggregated features to each snapshot, based on all tests/measurements prior to the timestamp of the snapshot:

- Most recent test/measurement,
- Mean of tests/measurements,
- Fitted slope of tests/measurements,
- Number of tests/measurements.

In total, 826,715 snapshots were created covering the set of patients in the timespan March 3rd, 2020 to January 5th, 2021.

For each patient snapshot, we defined *n*-day forecast prediction targets for ICU admission and use of mechanical ventilation. Each target described the *n*-day forecast for the patient, for instance whether the patient will be in hospital *n* days later.

By summing targets for snapshots with the same date, we obtained corresponding population wide *n*-day forecast *training targets*: The number of patients admitted to ICU *n* days from a given date, and the number of patients on mechanical ventilation *n* days from a given date. A limitation of constructing population wide *n*-day targets in this fashion is that they underestimate the ground truth due to late arriving patients; any *n*-day target cannot include patients added to the dataset in the *n* days after the prediction is made. This problem increases with *n*. To get a proper evaluation of our approach, we thus also consider the *true targets* for each training target above, by summing over the actual admitted and ventilated patients for each date.

## METHODS

### Evaluation setup

We applied *Random Forest (RF)*^9^ models for *n*-day forecasting of ICU admission and ventilator use for all patients with a positive SARS-CoV-2 test. We trained and evaluated the models in two different setups:

- **Monthly retraining:** To emulate the real-world use case in which models will be frequently retrained (possibly daily), we fitted models on all (training) data until the first of a month and evaluated them on data for the following month (test data), for each of the 7 last months of 2020. For instance, models trained on data until June 1^st^ were evaluated on data from June. Predictions from each month were then pooled together to be used for evaluation over the entire 7 months.
- **Second wave:** The dataset was split into the first and second wave (before and after June 1^st^). Models were fitted on the first wave (training data) and evaluated on the second wave (test data). Predictions from the entire second wave was used for evaluation in this setting.

For model fitting, missing BMI measurements were imputed based on the training data using *k*-nearest neighbor imputation considering only patients of the same sex (*k=100*). Missing in-hospital tests and measurements were set to 0.

### Model setup

We fitted random forests on the training set^9, 10^, with each individual decision tree in the ensemble trained on a bootstrap sample from the training set, leaving a set of *out-of-bag (OOB) samples* available for evaluating the performance of the RF^9^.

We used 500 trees and considered all features in each split, using the Gini-impurity as splitting criterion. We tuned the *maximum tree depth* (2, 4, 6 and 8) by performing grid search using the *Receiver Operating Characteristics Area Under the Curve (ROC-AUC)* computed on the OOB samples as evaluation metric.

Population wide (macro) forecasts were obtained from the individual (micro) prediction models by summation over the predicted probabilities for all individual patients for a given timestamp. Furthermore, we also computed the 95% confidence intervals (CI) by considering individual predictions as dichotomous Bernoulli variables and then summing their variances.

### Evaluation metrics

We evaluated the population wide forecast models by computing the *coefficient of determination (R*^2^) and the *maximum error (ME)* between the predictions and the true targets. Furthermore, we constructed plots of the true target and the forecast trajectory for both ICU admissions and ventilator use, including in the plots the training targets and the 95% CIs for the outputs.

We also evaluated the micro models for making individual predictions by computing the ROC-AUC scores over all test set predictions, as well as plotting the ROC curves.

We computed 95% confidence intervals for both R² and ROC-AUC scores by bootstrapping. Furthermore, the performance of the RF for predicting individual outcomes was compared to the baseline using the deLong test.^11^

### Baseline comparison models

We considered three baseline models:

- A standard *logistic regression (LogR)* micro-level model fitted to the individual patients and applied in the same fashion as the RF to make population wide forecasts. Continuous features were Z-normalized before the model was applied.
- A *static* micro-level model, which for each individual patient simply outputs the current state of the patient, that is, it predicts hospitalization if the patient is currently in hospital.
- A standard *linear regression (LinR)* macro-level model fitted directly on the true target curves (ICU admission or ventilated patients); for predicting the number of patients admitted to ICU or ventilated at day *D+n*, the model input was the number of patients at days *D, D-1, D-2, D-4, D-8,* and *D-12.* In contrast to our approach and the above baselines, this model was fitted on the true targets, which is possible because it works directly on the population level.

## RESULTS

### Demographics

Table 1 presents an overview of the data set, including the number of hospitalized, ICU admitted and ventilated patients. Figure 1 shows the distribution of snapshots over time. Patients were observed to change state multiple times during the observed time period for both ICU and ventilator use. Table 2 reports median, interquartile range (for continuous features) and percentage (for binary features) for age, sex, BMI, and comorbidities in each of the three data sets. The percentages of missing features are listed in Supplementary Table 1, including the percentages of missing features for the subset of patients in hospital.

**Figure 1:**
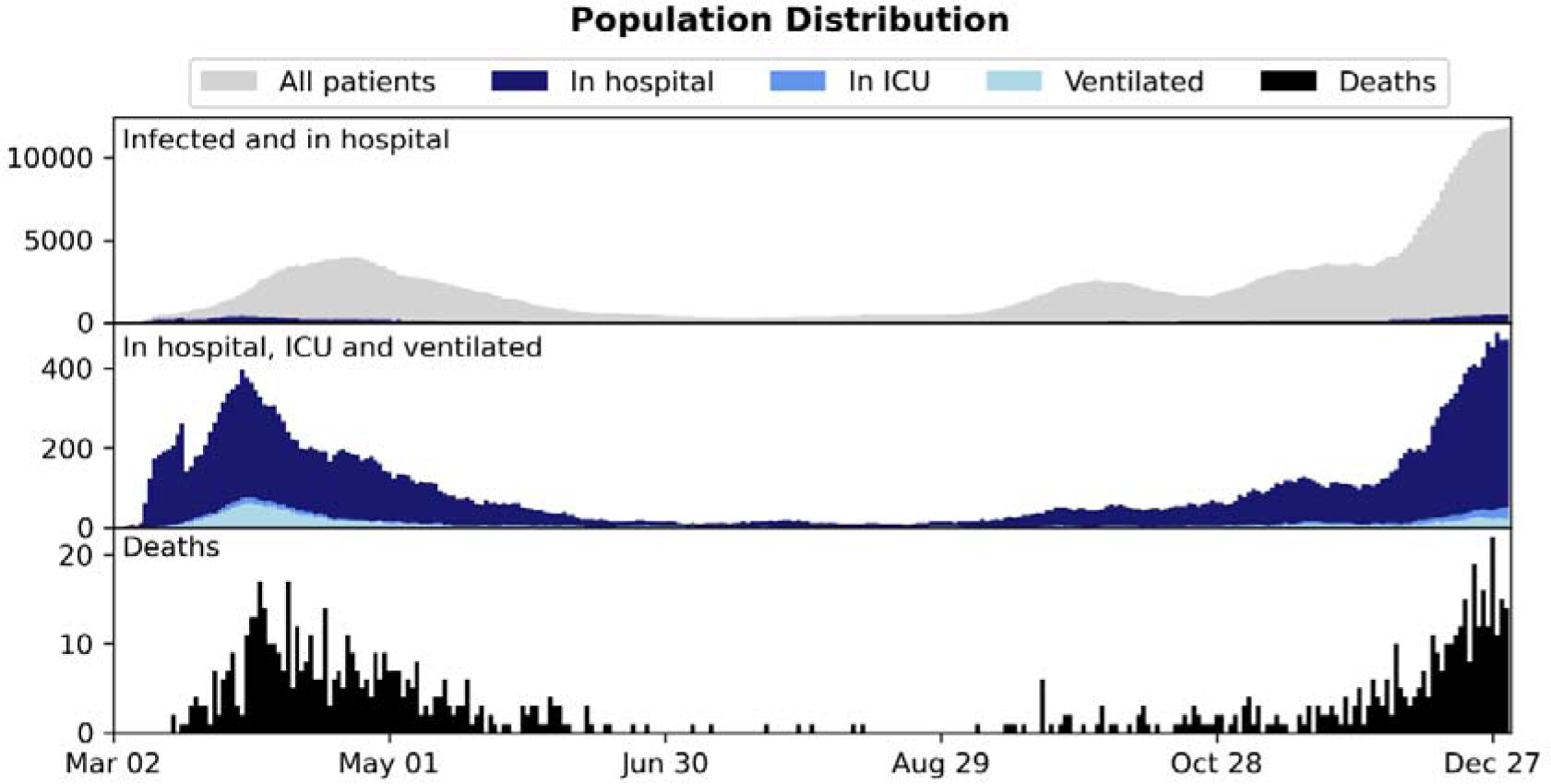
Distribution of patient 24-hour snapshots during the period consider in the study.

**Table 1:** An overview of the data sets and the number of patients in different states: patients tested positive with SARS-CoV-2, patients admitted to hospital, and patients admitted to ICU.

| Patient state | #Patients | #Snapshots |
| --- | --- | --- |
| Infected | 34,012 | 826,715 |
| In hospital | 5,028 | 120,245 |
| In ICU | 432 | 9,479 |

**Table 2:** Data set statistics: basic patient information (BMI, age, sex), patient comorbidities and smoker/non-smoker status. For binary variables, we report the occurrence in percent; for continuous variables, we report the median and interquartile range.

|  | Infected | Not admitted to hospital | Admitted to hospital | Not admitted to ICU | Admitted to ICU | Survivor | Non-survivor |
| --- | --- | --- | --- | --- | --- | --- | --- |
| # of patients | 34,012 | 28,984 | 5,028 | 4,596 | 432 | 33,113 | 899 |
| Age | 44 (27 - 60) | 40 (25 - 55) | 70 (53 - 80) | 70 (53 - 81) | 68 (58 - 75) | 43 (26 - 58) | 83 (75 - 89) |
| Body Mass Index | 24.8 (21.2 - 29.0) | 24.3 (20.5 - 28.4) | 26.2 (23.0 - 30.2) | 26.0 (22.9 - 30.1) | 27.8 (24.1 - 31.4) | 24.8 (21.2 - 29.0) | 24.5 (21.5 - 28.5) |
| Male | 43.5% | 41.9% | 53.3% | 51.7% | 70.4% | 43.3% | 52.7% |
| Diabetes | 6.1% | 2.6% | 26.3% | 21.7% | 75.2% | 5.4% | 30.9% |
| Ischemic heart disease | 0.7% | 0.0% | 4.9% | 4.5% | 9.5% | 0.6% | 7.0% |
| Heart failure | 0.0% | 0.0% | 0.2% | 0.2% | 0.0% | 0.0% | 0.2% |
| Arrhythmia | 1.0% | 0.0% | 6.9% | 6.4% | 11.3% | 0.7% | 13.2% |
| Stroke | 2.9% | 1.8% | 9.4% | 9.5% | 8.3% | 2.6% | 15.1% |
| Asthma | 5.7% | 3.4% | 18.5% | 18.3% | 20.8% | 5.2% | 21.2% |
| Arthritis | 0.6% | 0.4% | 1.6% | 1.6% | 1.6% | 0.6% | 2.2% |
| Osteoporosis | 1.9% | 1.2% | 6.4% | 6.6% | 4.2% | 1.7% | 11.8% |
| Dementia | 1.9% | 1.4% | 4.8% | 5.2% | 0.9% | 1.5% | 14.2% |
| Severe mental disorder | 0.8% | 0.7% | 1.7% | 1.7% | 1.9% | 0.8% | 2.0% |
| Immuno-deficiencies | 0.1% | 0.0% | 0.1% | 0.0% | 0.5% | 0.1% | 0.0% |
| Neurological manifestations | 7.9% | 6.5% | 15.7% | 16.0% | 12.7% | 7.4% | 24.9% |
| Cancer | 4.2% | 2.8% | 12.7% | 12.8% | 12.3% | 3.8% | 20.7% |
| Chronic kidney failure | 0.9% | 0.4% | 4.2% | 4.2% | 3.9% | 0.8% | 6.7% |
| Dialysis | 0.2% | 0.1% | 0.9% | 0.9% | 0.5% | 0.2% | 0.9% |
| Hypertension | 10.0% | 4.2% | 43.5% | 40.7% | 74.1% | 8.9% | 49.1% |
| Smoker | 17.0% | 17.4% | 15.8% | 16.3% | 11.3% | 17.0% | 16.3% |

### Monthly retrained

Table 3 reports the R^2^ and ME for the 5 and 10-day forecasts for the RF model. Figures 2 and 3 show the 5- and 10-day forecasts versus the training and the true targets respectively. The dates on the x-axes mark the retraining of models. The RF was found to perform very well when retrained monthly, obtaining an R² of 0.930 and 0.934 for 5-day forecasting of ICU admission and ventilator use, with performance dropping to 0.707 and 0.855 for 10-day forecasts.

**Figure 2:**
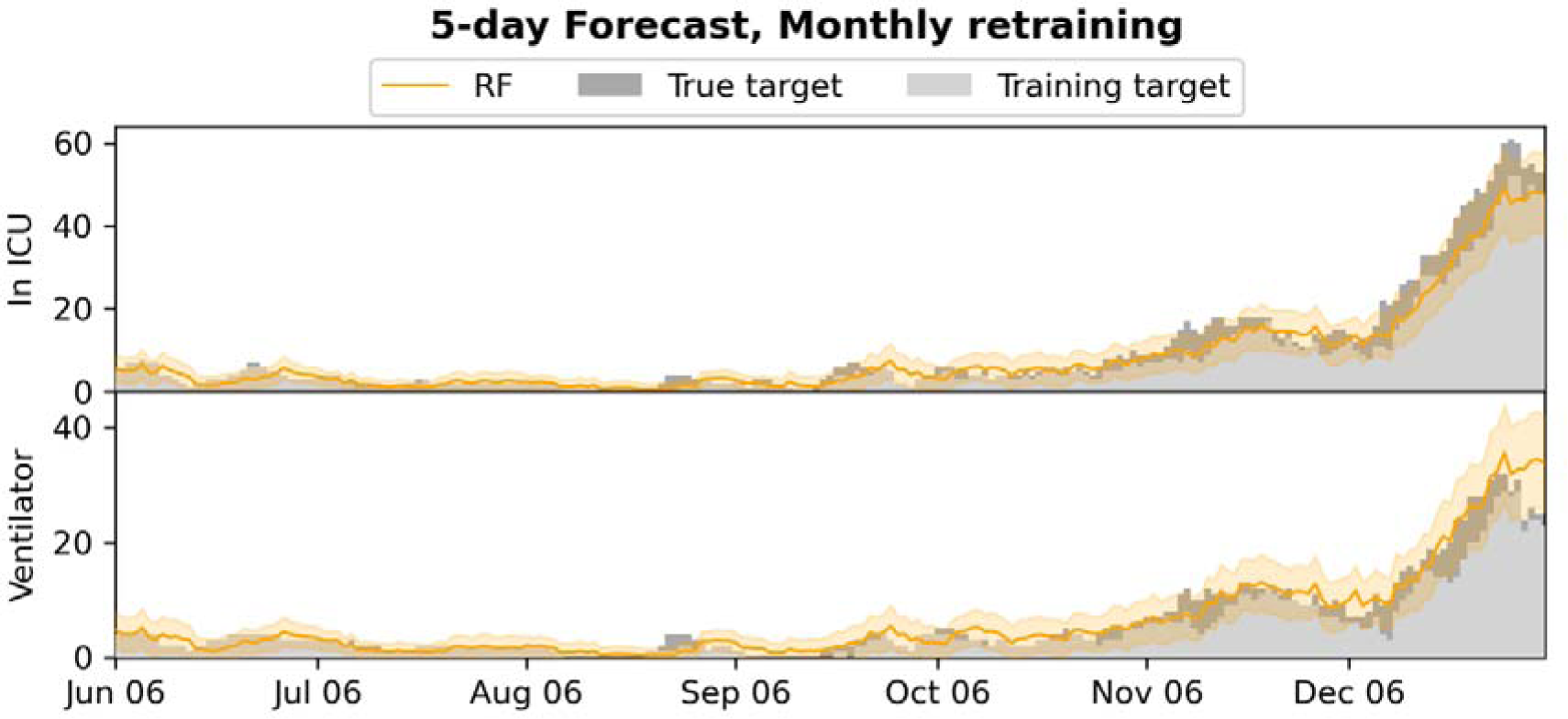
5-day forecasts of admission to ICU and use of mechanical ventilation, compared to the training and true targets in the monthly retrained setting. The estimated 95% confidence intervals are shown. The x-axis is marked with a date whenever a newly trained model is used.

**Figure 3:**
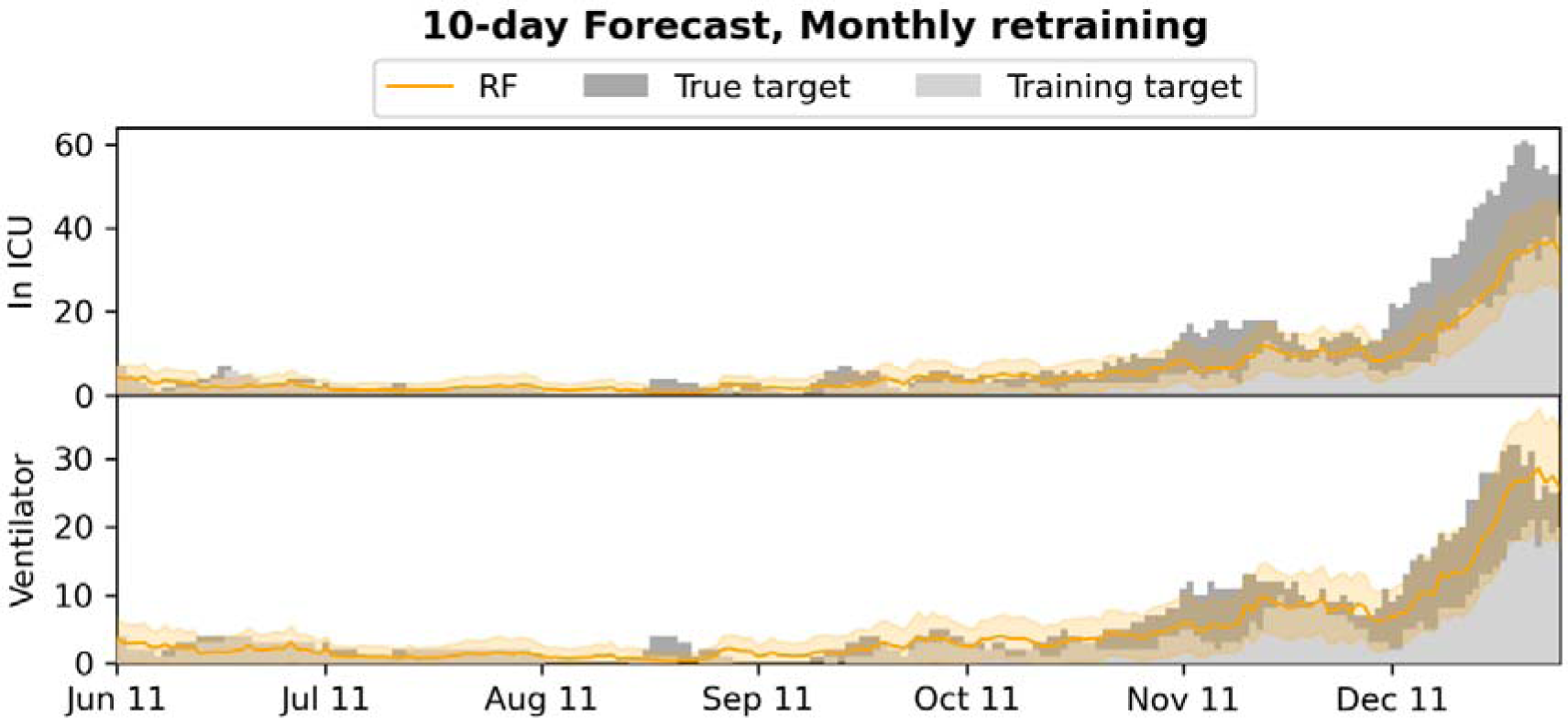
10-day forecasts of admission to ICU and use of mechanical ventilation, compared to the training and true targets in the monthly retrained setting. The estimated 95% confidence intervals are shown. The x-axis is marked with a date whenever a newly trained model is used.

**Table 3:** Results for 5- and 10-day forecasts for the monthly retrained setting. R^2^ and 95% confidence intervals (top), and ME (bottom) were computed on the pooled predictions for the last 7 months of 2020.

| n | ICU admission | Mechanical ventilation |
| --- | --- | --- |
| 5 | 0.930 (0.912 - 0.943) | 0.934 (0.900 - 0.958) |
|  | -15.6, 3.4 | -4.6, 9.5 |
| 10 | 0.707 (0.658 - 0.746) | 0.855 (0.812 - 0.889) |
|  | -27.1, 2.0 | -9.9, 4.5 |

Supplementary Figures 1 and 2 plot the baseline forecasts for the 5- and 10-day forecasts respectively. Supplementary Tables 2 and 3 present numerical results for the RF model and the baselines, including also results for hospital admission and mortality (with targets constructed in a similar fashion to the targets for ICU and ventilator). Compared to the baseline models, the RF performed better than all baselines for 5-day ICU admission and ventilator forecasts (without overlap of the 95% CIs). For 10-day ICU admission forecasts, the RF exhibited similar performance to the LinR model, and was outperformed by the LogR model.

### Second wave

Table 4 reports the R^2^ and ME for the 5 and 10-day forecasts for the RF model. Figures 4 and 5 show the 5- and 10-day forecasts versus the training and true targets for both the first wave (training data) and the second wave (test data). The model was found to generalize well to the second wave for both 5- and 10-day ICU admission and ventilator forecasts, obtaining ROC-AUCs between 0.835 and 0.958.

**Figure 4:**
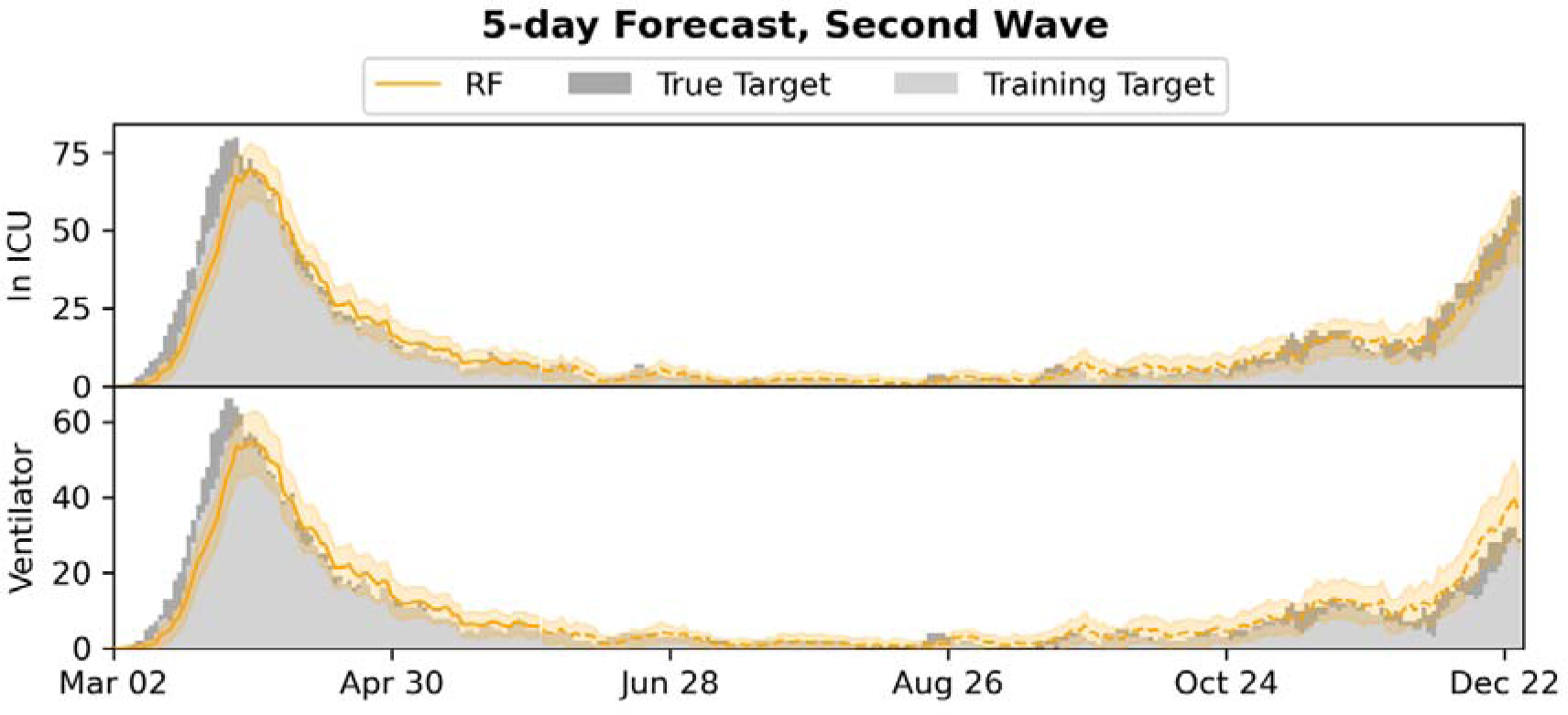
5-day forecasts of admission to ICU and use of mechanical ventilation, compared to the training and true targets in the second wave setting. Predictions and targets for both the first wave (training data) and the second wave (test data, dashed) are shown, as well as the 95% confidence intervals.

**Figure 5:**
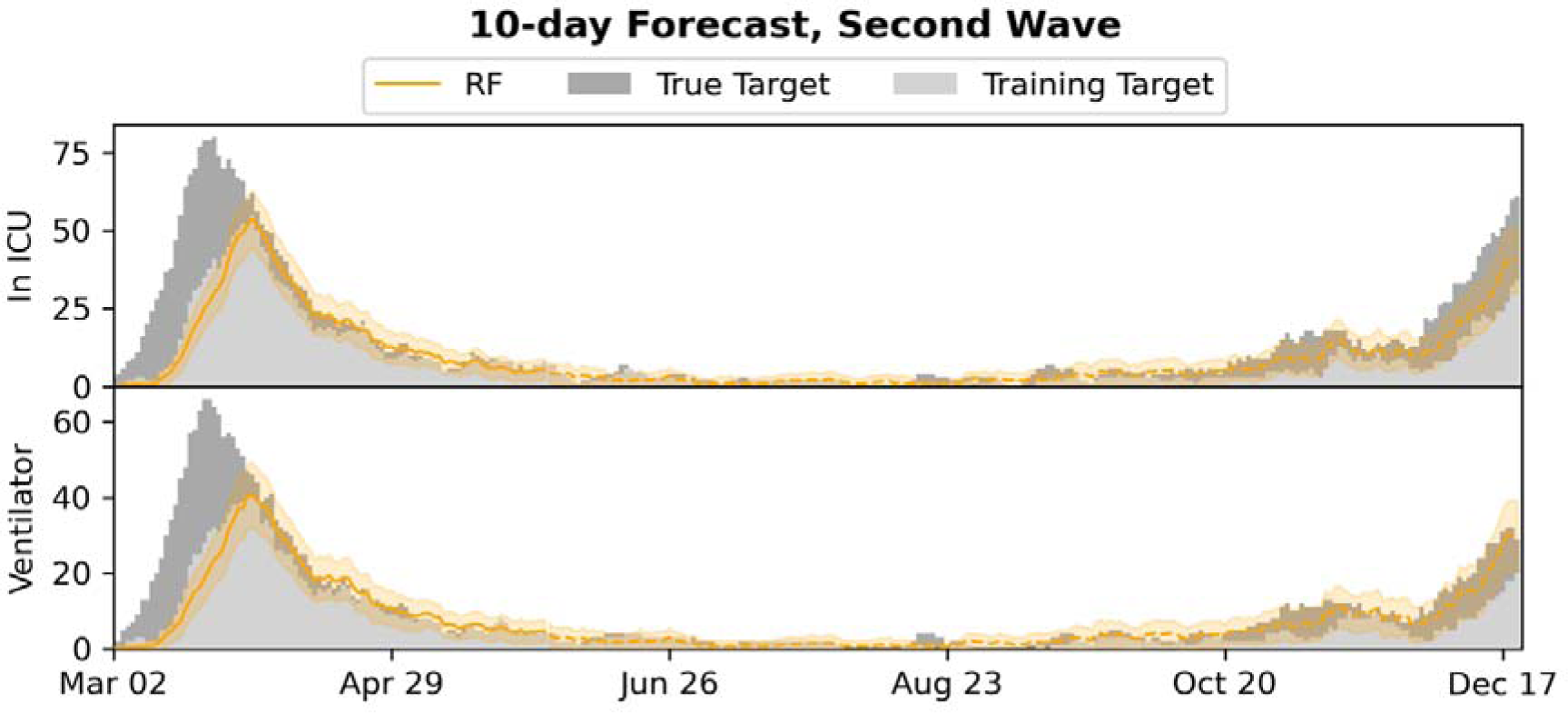
10-day forecasts of admission to ICU and use of mechanical ventilation, compared to the training and true targets in the second wave setting. Predictions and targets for both the first wave (training data) and the second wave (test data, dashed) are shown, as well as the 95% confidence intervals.

**Table 4:** Results for 5- and 10-day forecasts for the second wave setting. R^2^ and 95% confidence intervals (top), and ME (bottom) were computed on the predictions for the last 7 months of 2020.

| n | ICU admission | Mechanical ventilation |
| --- | --- | --- |
| 5 | 0.958 (0.945 - 0.968) | 0.866 (0.805 - 0.915) |
|  | -12.3, 3.9 | -4.0, 14.6 |
| 10 | 0.835 (0.796 - 0.865) | 0.900 (0.863 - 0.924) |
|  | -20.9, 2.2 | -8.2, 7.6 |

Supplementary Figures 3 and 4 plot the baseline forecasts for the 5- and 10-day forecasts respectively. Supplementary Tables 4 and 5 present numerical results for the RF model and the baselines, including results for hospital admission and mortality. The RF outperforms all baselines for 5- and 10-day ICU admission and ventilator forecasts; in particular it outperforms the population level LinR model (without overlap of the 95% CIs).

### Individual forecasts

Supplementary Tables 6 and 7 report ROC-AUC for the individual 5- and 10-day predictions for the macro models RF and LogR when retrained monthly, including results for prediction of hospital admission and mortality. Supplementary Figures 5 and 6 show the corresponding ROC curves. For all 5- and 10-day forecasting targets, the RF was found to obtain ROC-AUC scores above 0.95 and was found to perform significantly better than LogR (*p* < 0.0001) by application of the deLong test.

## DISCUSSION

In this study, we demonstrate how COVID-19 micro-predictions made at the level of the individual SARS-CoV-2 positive patient, can be extrapolated to a population-wide macro- prediction by modelling incremental patient data as this becomes available in the EHR system. Based on evaluation in a cohort with 34,012 SARS-CoV-2 positive patients, we find that, using an RF, accurate 5-day and 10-day forecasting of ICU admissions and ventilator use can be achieved using initial datapoints including age, sex, BMI and comorbidities with additional datapoints including lab tests and vital parameters added as these become available. As expected, the results for the 10-day forecasts were in general less precise compared to the 5-day predictions. When using regular retraining, the RF approach outperformed all baseline methods for 5-day forecasts and was only significantly outperformed by logistic regression for 10-day ICU admission forecasts, as explained below. The setting with regular retraining best resembles the expected real-life deployment, during which the model will be retrained on the latest available data even more frequently. When only a single model trained on the first wave was used, the RF still generalized well for forecasting admission to ICU and use of ventilation, beating every baseline for both 5- and 10-day forecasts, including the macro-level LinR model, confirming that, in this case, the micro-to-macro approach handles shifts in the distribution (on the macro/population level) better.

While we reported results for only monthly retraining, the data processing and RF implementation is so efficient that the models can be retrained on the entire cohort daily as new data arrives, allowing for real-time deployment of the most up-to-date models.

The system is currently limited by missing access to early SARS-CoV-2 tests, which means patients are not included in the input early, leading to the model producing underestimates of the true admission/ventilation numbers. This can be seen by inspection of the plots of the forecast trajectories and training/true targets (e.g., Figures 2 and 3). The model consistently underestimates the true targets due to the training targets being underestimates themselves. The problem increases with the number of days *n* we look ahead in the forecasts, as can be seen by comparing the performance of the 5- and 10-day models. We also evaluated the model for prediction of hospital admissions, but since many patients are not confirmed positive until after admission, the issue was even more pronounced, and the model severely underestimated the true admission numbers (see Supplementary Tables 2 and 3), even though the model performed well for predicting for individual patients.

However, our focus is on ICU admission and use of ventilation, because constraints on ICU and ventilators are one of the main issues in capacity planning in hospitals ^12^– and for the 5- and 10-day forecasts of ICU admission and use of ventilation, the training targets and the true targets were fairly close, leading to good model performance and useable forecasts, with true targets often within the 95% CI.

When looking at the prediction accuracy on individual level, LogR performed worse than RF. In our experiments, the logistic regression had a tendency to overestimate the individual risks, that is, the training targets. Because for 10-day forecasts the training targets underestimate the true values, LogR gave better results on the true values for 10-days ICU prediction despite being less accurate on the individual level.

The problem that the training targets underestimate the true targets may be reduced by access to earlier SARS-CoV-2 testing. While the Danish test capacity during the second wave was improved and patients in general have been tested early (before hospital admission), we only have access to tests performed by the regional test system and not by private vendors or the national test centers. Thus, we only have access to a limited number of positive tests before admission and for many of the patients we only have a confirmatory in hospital test, leading to a very short time between test and hospital admission, and thus the discrepancy between the targets and actual number of ICU admitted or ventilated patients.

We expect the accuracy of the micro-to-macro approach to increase with access to these early tests, which might even make hospital admission forecasting feasible, however this will rarely be possible due to the number of different providers of COVID-19 tests. A more feasible alternative would be adjustment of predictions by use of macro model predictions, for instance by estimation of the total number of infected expected to arrive within the *n*-day forecasting window.

The findings mirror previously published results from the United Kingdom COVID-19 Capacity Planning and Analysis System (CPAS)^13^. In contrast to the CPAS system, our model captures patients at the time of their positive SARS-CoV-2 test result, with the ability to extract EHR information for all patients irrespective of hospital admission status. Even when we lack access to the early tests for many patients, this allows us to forecast ICU admission and ventilator use, even for non-hospitalized patients. This approach is feasible due to data sharing between the EHR system and Danish national registries, where detailed information on diagnoses codes, previous hospital admissions and pharmaceutical use is available for all Danish citizens. As such, the system can model basic demographic and healthcare datapoints even for non-hospitalized patients, which aids the prediction of need for hospital admission on a population-wide scale. However, as mentioned, the approach is limited by patients with late positive test results. In practice, the hospitalization model has to be adjusted, for instance by extending the model by modelling COVID-19 spread as done by CPAS and then adjusting the hospitalization model accordingly. Even better would be access to the early Danish PCR tests performed outside of the hospitals, which we hypothesize will alleviate the problem.

Our approach differs from conventional epidemiological analyses, where modelling is based on the number of patients tested positive, calculated infection rates and social mobility models for a specific infection during a pandemic with subsequent forecasting^14^. The generalization ability and performance of the RF model between the 1^st^ and 2^nd^ COVID-19 waves of 2020 shows the potential of modelling risk predictions at the individual level based on EHR data with subsequent extrapolation to population wide modelling in this as well as future pandemics. Furthermore, the approach could be ported to other disease settings, including a hospital-wide ICU admission forecasting, provided relevant disease modalities could be modelled by dedicated ML systems.

Pivotal for the success of this approach is, however, an ability to rapidly mobilize EHR data for the purpose of ML modelling, thus indicating the need for maintaining updated data transfer protocols from the EHR system to a High-Performance Computing (HPC) platform capable of training ML models in a secure environment. Such EHR and HPC integration should ideally be maintained even between pandemic surges, owing to the often-rapid spread and unknown features of pandemics.

Furthermore, accumulating data indicates that the patient characteristics of the COVID-19 pandemic change from the first to subsequent waves. Indeed, reports have indicated that the current 2^nd^ COVID-19 pandemic wave is characterized by an altered disease spectrum and updated treatment protocols^15^, resulting in potentially milder disease trajectories with lower ICU and ventilator requirements ^3^. As such, effective forecasting models need to be robust towards demographic fluctuations, which is exemplified in our approach, where the predictive performances of the models were retained for both 1^st^ and 2^nd^ wave patients in this study. Furthermore, these fluctuations can be addressed by frequent model retraining, as evidenced by the improved performance of the monthly retraining method demonstrated here. While demographics may change, the pandemic progression carry the inherent risk of mutations in the viral genome altering the disease severity, as is seen SARS-CoV-2 variations such as the D614G and B117 currently on the rise in multiple countries and is expected to give rise to a 3^rd^ wave in Denmark^16, 17^. As such, predictive models could ideally be dynamically retrained on emerging data from multiple data sources, including potentials such as viral sequencing data in combination with EHR data.

Furthermore, adding additional data feeds (e.g., imaging data) to hybrid prediction models could potentially augment the value of such systems, which has been suggested by previous studies^18–20^. The caveat is, however, that such systems require integration of healthcare and population specific data, enabling extraction of EHR data on a community wide rather than a hospital specific scale. Healthcare systems operating as individual units rather than covering the entirety of a regional population, may thus not be optimally suited for real-time deployment of these prediction models.

This study has several limitations. First, we model data retrospectively and have not demonstrated a prospective value in this study. As such, although accurate on retrospective data predictions, novel features of a potential third COVID-19 wave could affect model performance. Furthermore, we model a selected subset of patient-derived variables based on previous experience^7^, although other data points could affect the model’s classification ability.

Secondly, we have not performed external validation on a separate cohort. This, however, may not be desirable due to several factors: The model should be trained to forecast locally, and transferring to different healthcare systems with different social and geographical factors would require retraining of the model to capture these effects. We have also recently demonstrated that transferring COVID-19 ML prediction models between health care systems internationally results in a reduction of the classification precision, presumably due to inherent differences between healthcare systems even though patient demographics are comparable^7^.

Thirdly, as mentioned, the model could benefit from either adjustment based on macro predictions, or access to earlier SARS-CoV-2 tests for Danish patients, before deployment. This might also make hospital admission feasible.

In conclusion, this study demonstrates that transferring ML based patient level predictions for COVID-19 to a population wide-scale for the purpose of 5-day and 10-day resource use predictions, is feasible and should be considered for the current and future pandemic events.

## Data Availability

Due to patient confidentiality issues, the data cannot legally be made publicly available without authorisation from the approving legal bodies.

## Contributors

Stephan Sloth Lorenzen, Mads Nielsen, Espen Jimenez-Solem, Tonny Studsgaard Petersen, Anders Perner, Hans-Christian Thorsen-Meyer, Christian Igel and Martin Sillesen. SSL, MN, EJS, TSP, AP, HCTM, CI and MS participated in study conception. SSL, MN and CI performed model construction and validation. SSL and MS drafted the original manuscript which was critically reviewed by all authors. All authors have read and approved the final manuscript.

## Declaration of interests

We declare no competing interests

## Data Availability Statement

Data used for the purpose of this manuscript cannot be publicly shared due to patient confidentiality issues.

## Acknowledgements

We gratefully acknowledge the support of the Novo Nordisk Foundation through grants #NNF20SA0062879 and #NNF19OC0055183 to MS and #NNF20SA0062879 to MN.

The foundation took no part in project design, data handling and manuscript preparation

## SUPPLEMENTARY FIGURES

**Supplementary Figure 1:**
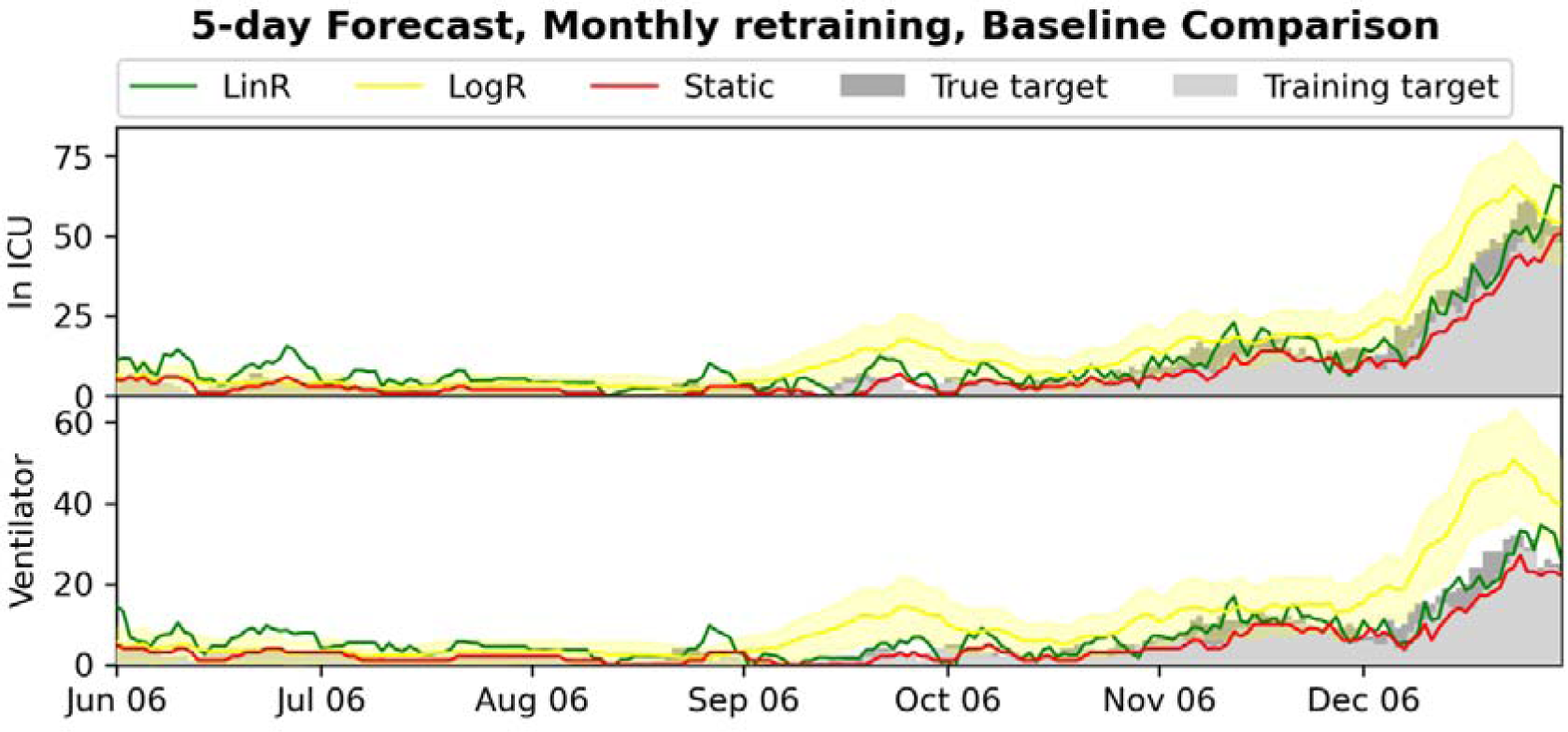
Plot of the 5-day forecasts of admission to ICU and use of mechanical ventilation for the different baselines in the monthly retrained setting. Training and true targets are shown, as well as estimated 95% confidence intervals for LogR (note that LinR and static do not admit a 95% CI).

**Supplementary Figure 2:**
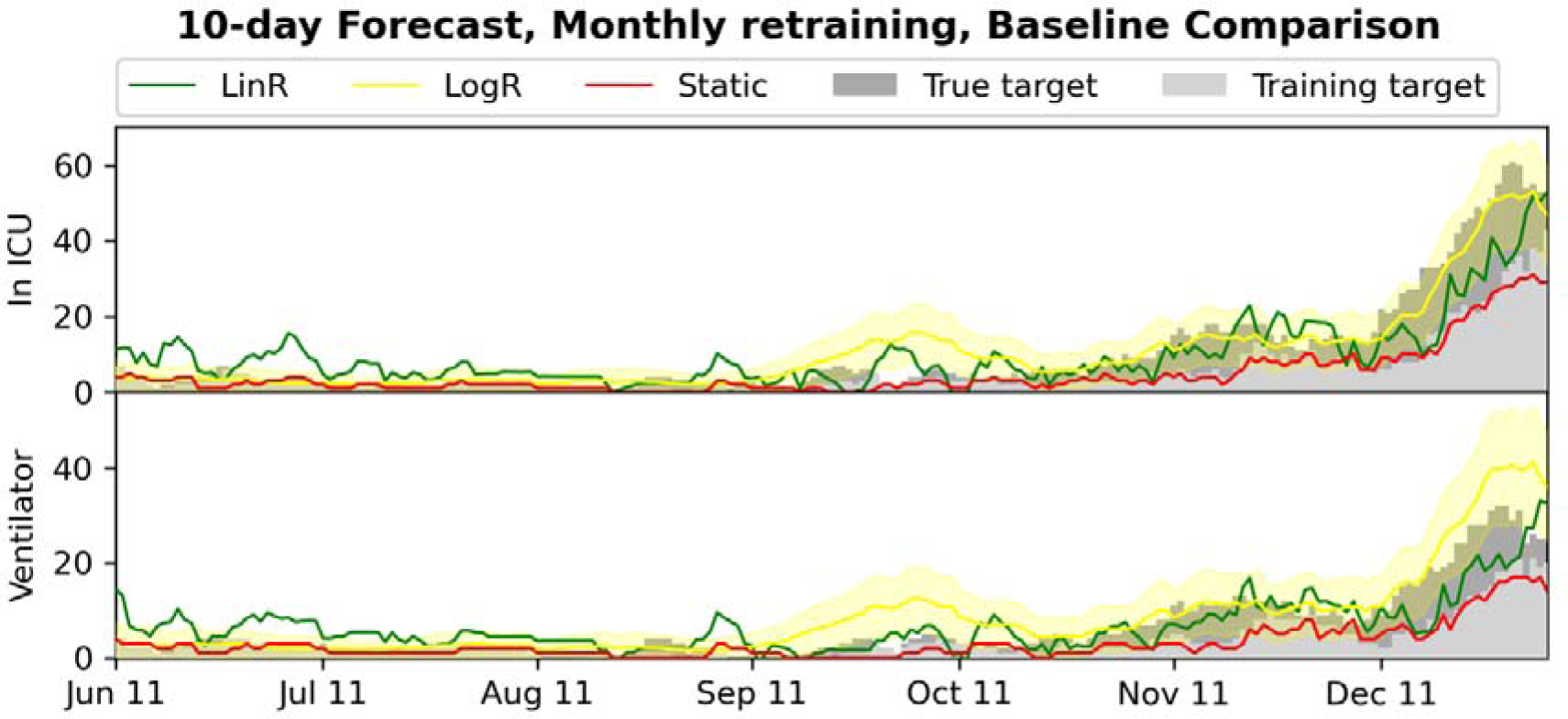
Plot of the 10-day forecasts of admission to ICU and use of mechanical ventilation for the different baselines in the monthly retrained setting. Training and true targets are shown, as well as estimated 95% confidence intervals for LogR.

**Supplementary Figure 3:**
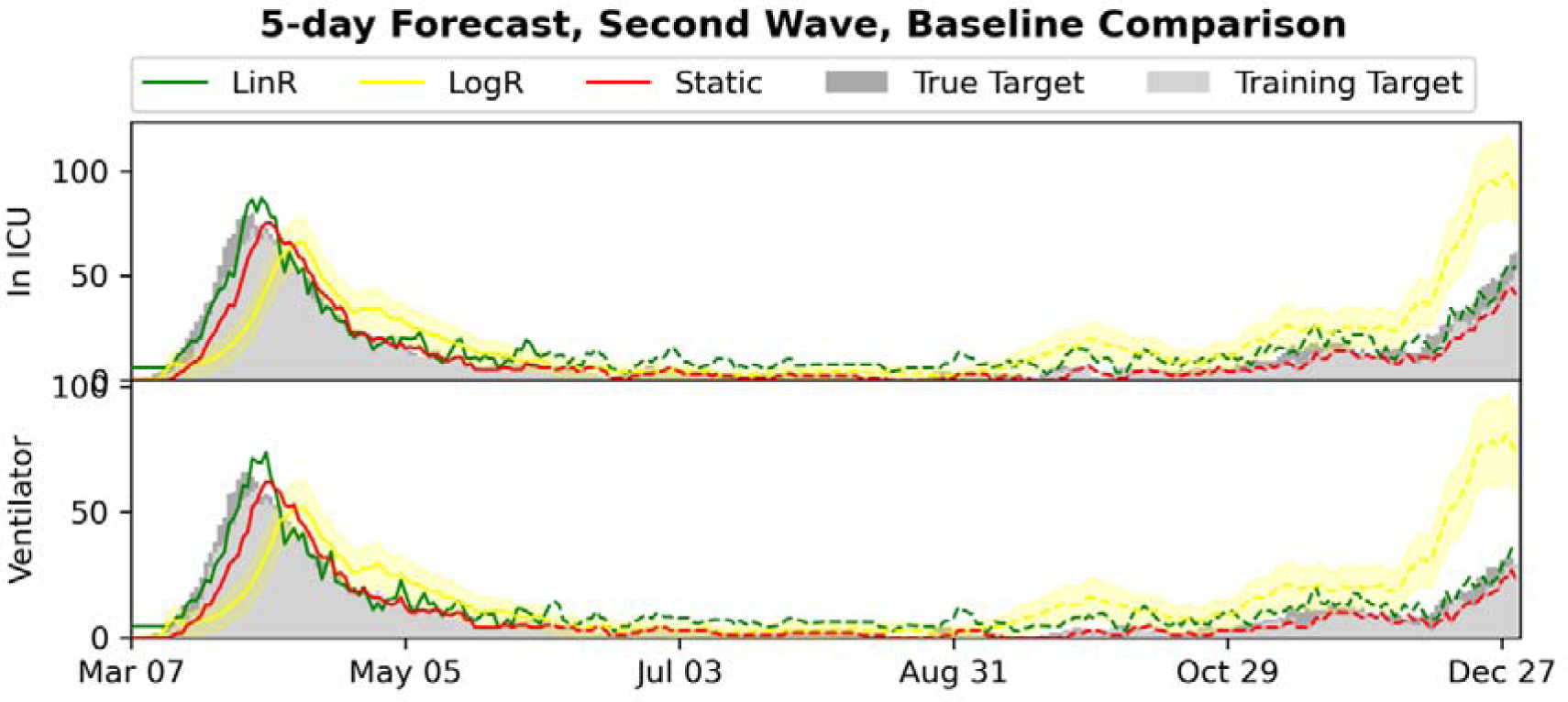
Plot of the 5-day forecasts of admission to ICU and use of mechanical ventilation for the different baselines in the second wave setting. Predictions, training and true targets are shown for both the first wave (training data) and the second wave (test data), as well as estimated 95% confidence intervals for LogR.

**Supplementary Figure 4:**
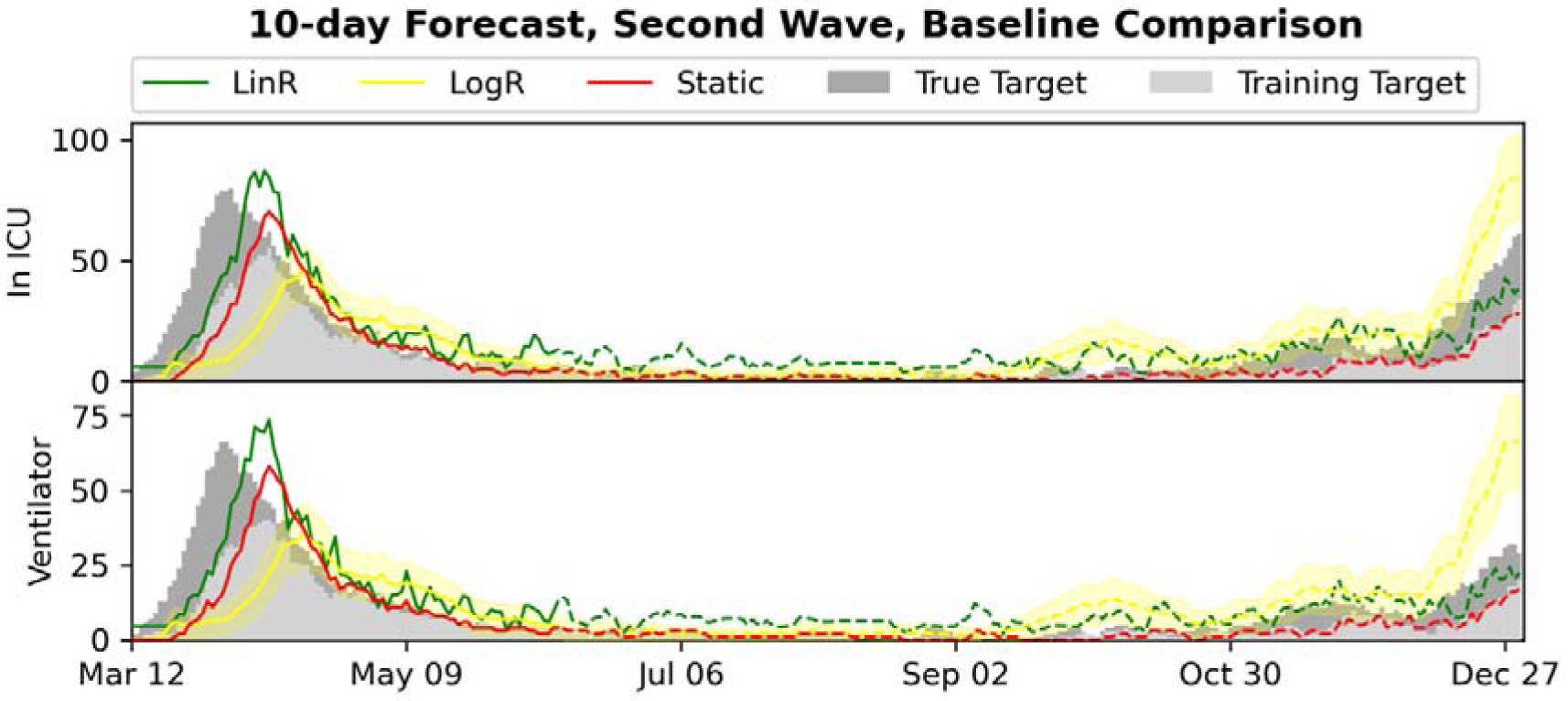
Plot of the 10-day forecasts of admission to ICU and use of mechanical ventilation for the different baselines in the second wave setting. Predictions, training and true targets are shown for both the first wave (training data) and the second wave (test data), as well as estimated 95% confidence intervals for LogR.

**Supplementary Figure 5:**
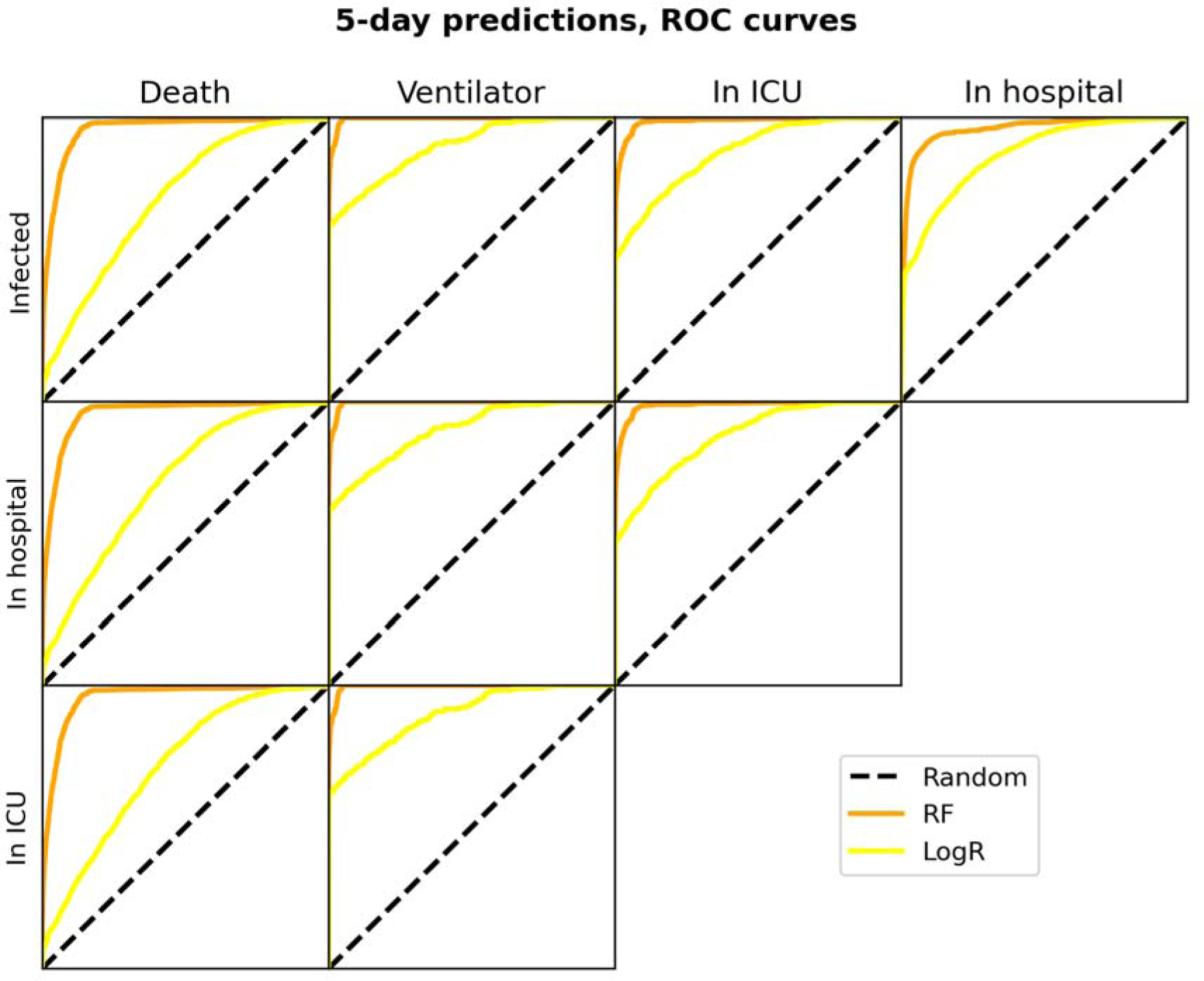
ROC curves for evaluation of the individual 5-day RF and LogR predictions in the monthly retrained setting.

**Supplementary Figure 6:**
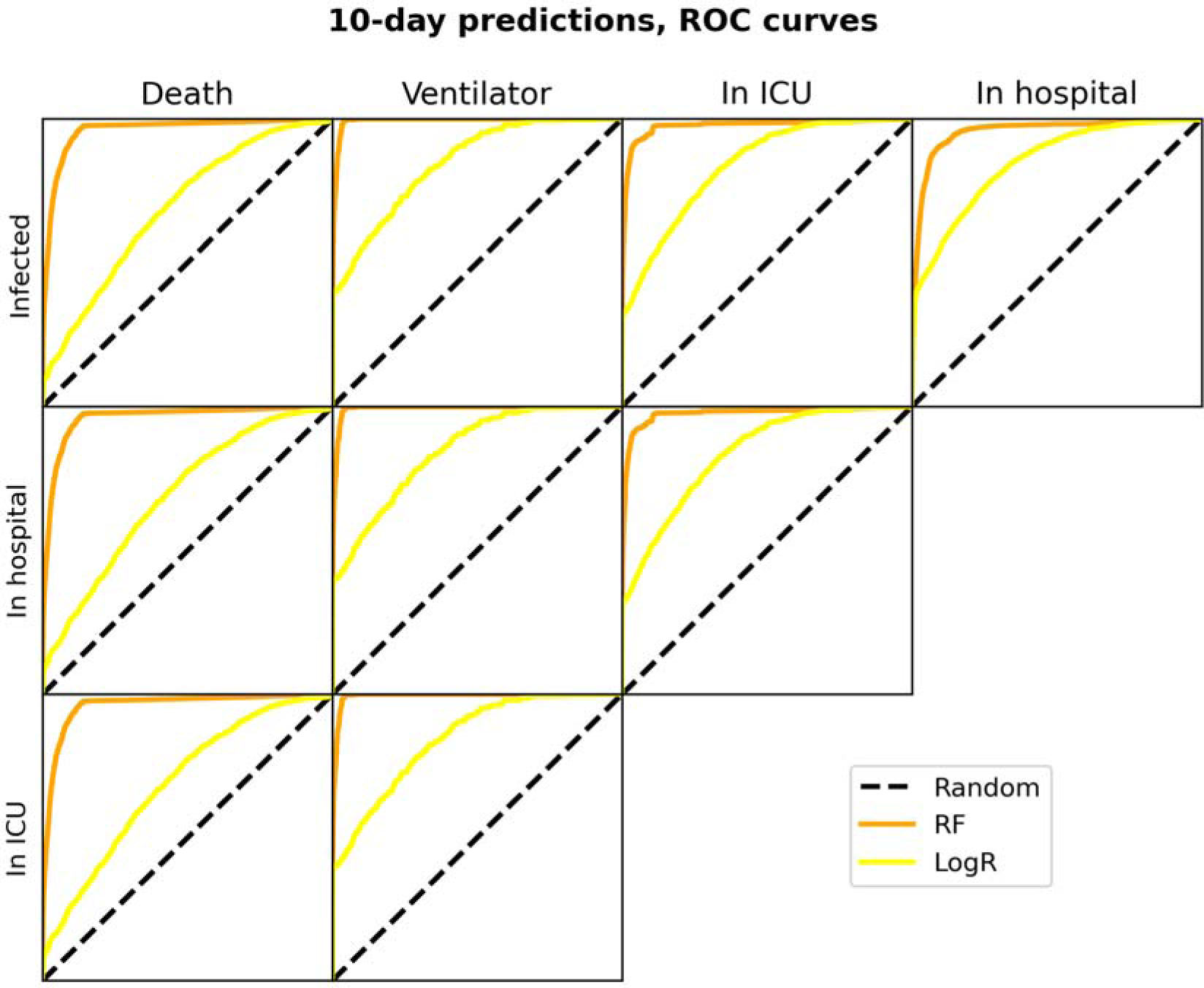
ROC curves for evaluation of the individual 5-day RF and LogR predictions in the monthly retrained setting.

## SUPPLEMENTARY TABLES

**Supplementary Table 1:**
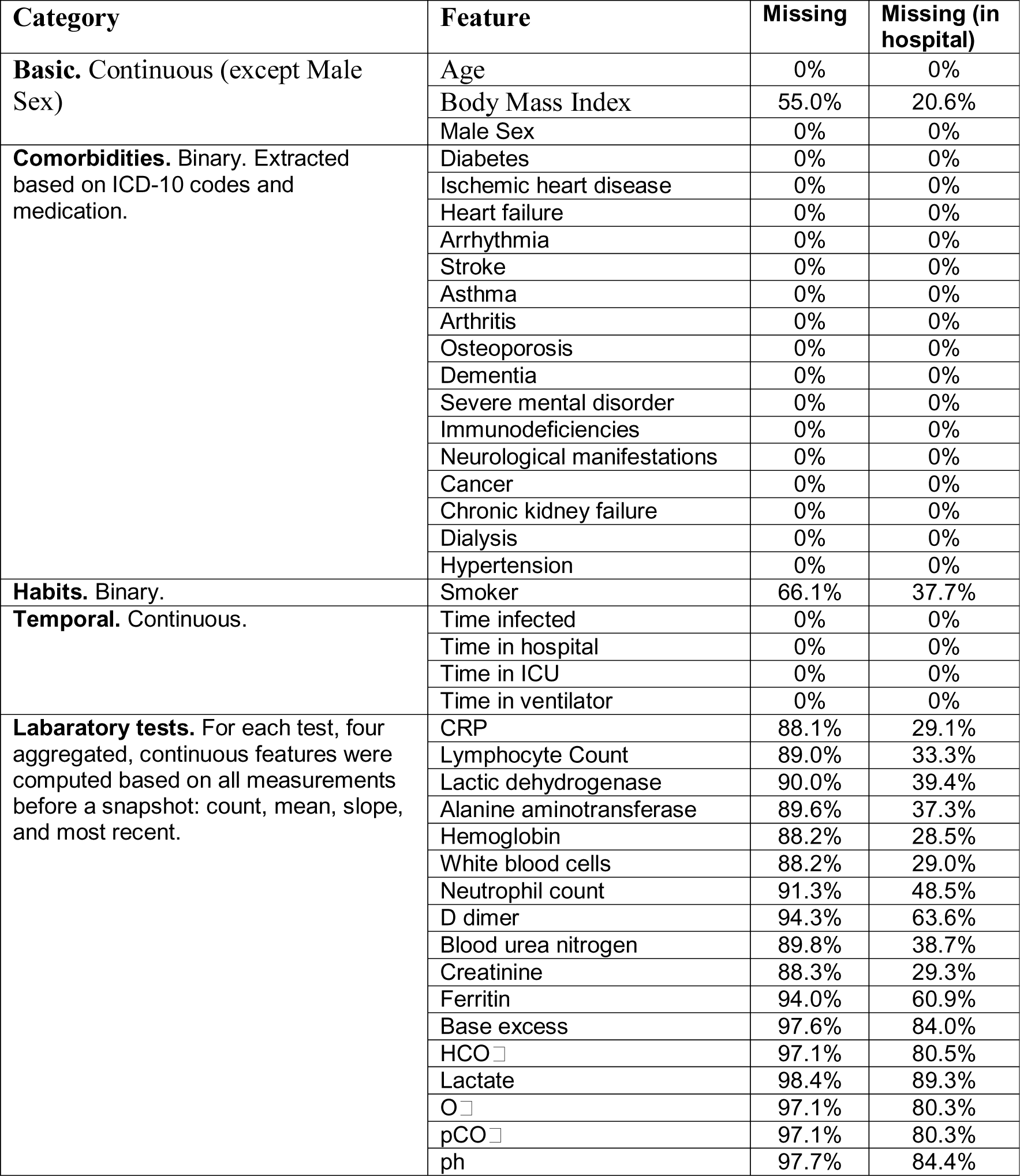
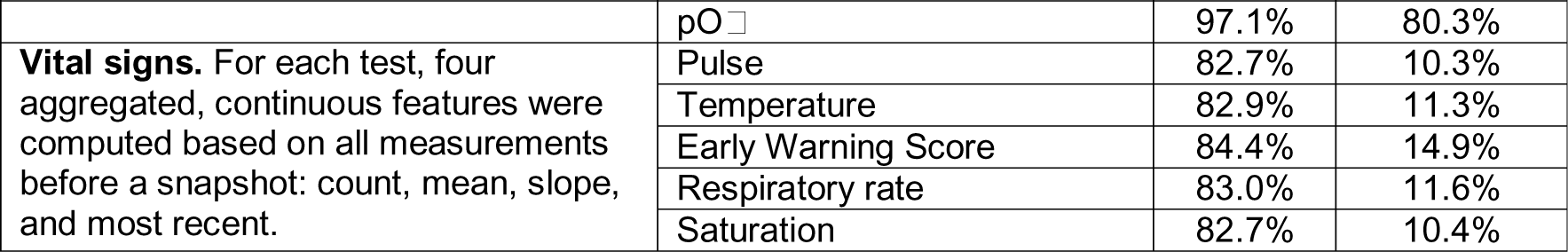
Feature categories, information, feature names and percentage missing features. For missing features, the percentages of missing features for all infected patients and the subset consisting of hospitalized patients are reported.

**Supplementary Table 2:** 5-day forecasting results for the RF and baseline models in the monthly retrained setting. R^2^ and 95% CIs (top), and ME (bottom) were computed on the pooled predictions for the last 7 months of 2020.

| Model | Hospital admission | ICU admission | Mechanical ventilation | Mortality |
| --- | --- | --- | --- | --- |
| RF | 0.714<br>(0.669 - 0.749) | 0.930<br>(0.911 - 0.944) | 0.934<br>(0.9 - 0.959) | 0.920<br>(0.885 - 0.946) |
|  | -229.0, 3.8 | -15.6, 3.4 | -4.6, 9.5 | -17.2, 27.5 |
| LogR | 0.861<br>(0.844 - 0.878) | 0.837<br>(0.761 - 0.888) | 0.055<br>(-0.177 - 0.203) | 0.791<br>(0.662 - 0.864) |
|  | -185.9, 41.3 | -2.8, 15.9 | -2.2, 21.7 | -6.5, 22.5 |
| LinR | 0.910<br>(0.872 - 0.934) | 0.866<br>(0.804 - 0.898) | 0.778<br>(0.677 - 0.836) | - |
|  | -147.8, 64.0 | -13.5, 12.8 | -9.3, 10.5 | - |
| Static | 0.883<br>(0.847 - 0.912) | 0.853<br>(0.811 - 0.882) | 0.819<br>(0.766 - 0.855) | - |
|  | -150.0, 6.0 | -20.0, 3.0 | -11.0, 2.0 | - |

**Supplementary Table 3:** 10-day forecasting results for the RF and baseline models in the monthly retrained setting. R^2^ and 95% CIs (top), and ME (bottom) were computed on the pooled predictions for the last 7 months of 2020.

| Model | Hospital admission | ICU admission | Mechanical ventilation | Mortality |
| --- | --- | --- | --- | --- |
| RF | 0.214<br>(0.144 - 0.268) | 0.707<br>(0.660 - 0.749) | 0.855<br>(0.809 - 0.890) | 0.863<br>(0.838 - 0.886) |
|  | -376.1, 1.6 | -27.1, 2.0 | -9.9, 4.5 | -50.2, 9.6 |
| LogR | 0.404<br>(0.341 - 0.452) | 0.916<br>(0.868 - 0.942) | 0.716<br>(0.622 - 0.787) | 0.856<br>(0.762 - 0.906) |
|  | -321.4, 3.4 | -11.7, 12.4 | -2.9, 15.8 | -22.5, 33.8 |
| LinR | 0.790<br>(0.725 - 0.838) | 0.738<br>(0.655 - 0.796) | 0.641<br>(0.522 - 0.709) | - |
|  | -239.8, 66.3 | -26.5, 11.6 | -13.5, 10.5 | - |
| Static | 0.679<br>(0.622 - 0.726) | 0.539<br>(0.462 - 0.588) | 0.468<br>(0.365 - 0.540) | - |
|  | -251.0, 10.0 | -33.0, 2.0 | -17.0, 2.0 | - |

**Supplementary Table 4:** 5-day forecasting results for the RF and baseline models in the second wave setting. R^2^ and 95% CIs (top), and ME (bottom) were computed on the predictions for the second wave (last 7 months of 2020).

| Model | Hospital admission | ICU admission | Mechanical ventilation | Mortality |
| --- | --- | --- | --- | --- |
| RF | 0.760<br>(0.730 - 0.783) | 0.958<br>(0.944 - 0.968) | 0.866<br>(0.803 - 0.916) | 0.836<br>(0.720 - 0.912) |
|  | -212.1, 4.0 | -12.3, 3.9 | -4.0, 14.6 | -5.6, 46.7 |
| LogR | 0.979<br>(0.972 - 0.984) | -0.116<br>(-0.475 - 0.133) | -3.728<br>(-4.594 - -3.045) | -0.243<br>(-0.575 - -0.051) |
|  | -81.4, 48.8 | -2.8, 48.9 | -2.1, 52.7 | -4.3, 58.8 |
| LinR | 0.819<br>(0.747 - 0.857) | 0.802<br>(0.699 - 0.853) | 0.603<br>(0.408 - 0.707) | - |
|  | -136.0, 67.5 | -10.7, 13.1 | -7.6, 11.2 | - |
| Static | 0.883<br>(0.847 - 0.911) | 0.853<br>(0.812 - 0.883) | 0.819<br>(0.760 - 0.858) | - |
|  | -150.0, 6.0 | -20.0, 3.0 | -11.0, 2.0 | - |

**Supplementary Table 5:**
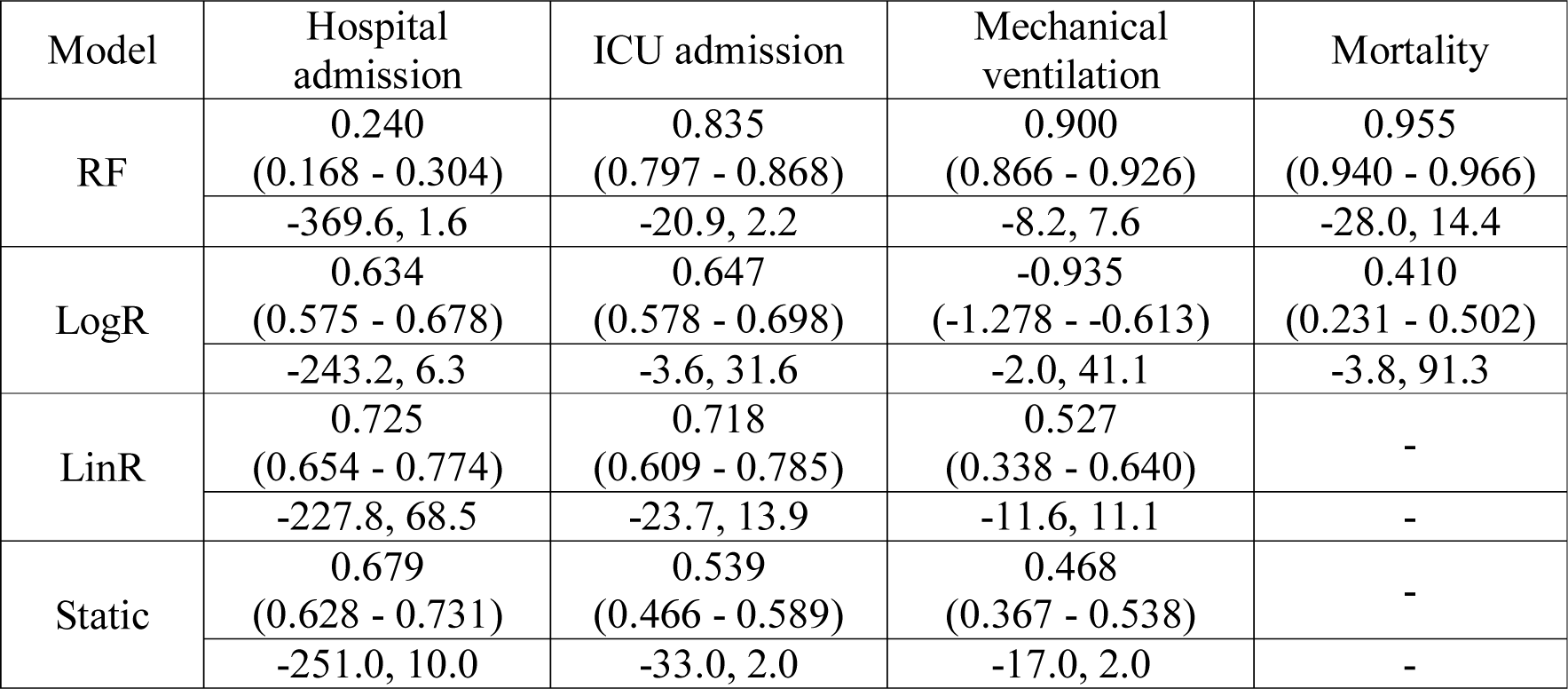
10-day forecasting results for the RF and baseline models in the second wave setting. R^2^ and 95% CIs (top), and ME (bottom) were computed on the predictions for the second wave (last 7 months of 2020).

| Model | Hospital admission | ICU admission | Mechanical ventilation | Mortality |
| --- | --- | --- | --- | --- |
| RF | 0.240<br>(0.168 - 0.304) | 0.835<br>(0.797 - 0.868) | 0.900<br>(0.866 - 0.926) | 0.955<br>(0.940 - 0.966) |
|  | -369.6, 1.6 | -20.9, 2.2 | -8.2, 7.6 | -28.0, 14.4 |
| LogR | 0.634<br>(0.575 - 0.678) | 0.647<br>(0.578 - 0.698) | -0.935<br>(-1.278 - -0.613) | 0.410<br>(0.231 - 0.502) |
|  | -243.2, 6.3 | -3.6, 31.6 | -2.0, 41.1 | -3.8, 91.3 |
| LinR | 0.725<br>(0.654 - 0.774) | 0.718<br>(0.609 - 0.785) | 0.527<br>(0.338 - 0.640) | - |
|  | -227.8, 68.5 | -23.7, 13.9 | -11.6, 11.1 | - |
| Static | 0.679<br>(0.628 - 0.731) | 0.539<br>(0.466 - 0.589) | 0.468<br>(0.367 - 0.538) | - |
|  | -251.0, 10.0 | -33.0, 2.0 | -17.0, 2.0 | - |

**Supplementary Table 6:** ROC-AUC score and 95% confidence intervals for the RF and LogR models used for making 5-day individual predictions in the monthly retrained setting. For every target, RF was found to perform significantly better than LogR when tested with the DeLong test (*p* < 0.0001).

| Model | Hospital admission | ICU admission | Mechanical ventilation | Mortality |
| --- | --- | --- | --- | --- |
| RF | 0.960<br>(0.957 - 0.962) | 0.986<br>(0.982 - 0.990) | 0.995<br>(0.994 - 0.996) | 0.956<br>(0.952 - 0.961) |
| LogR | 0.870<br>(0.865 - 0.875) | 0.879<br>(0.865 - 0.892) | 0.898<br>(0.882 - 0.915) | 0.716<br>(0.705 - 0.727) |

**Supplementary Table 7:** ROC-AUC score and 95% confidence intervals for the RF and LogR models used for making 10-day individual predictions in the monthly retrained setting. For every target, RF was found to perform significantly better than LogR when tested with the DeLong test (*p* < 0.0001).

| Model | Hospital admission | ICU admission | Mechanical ventilation | Mortality |
| --- | --- | --- | --- | --- |
| RF | 0.954<br>(0.950 - 0.958) | 0.975<br>(0.967 - 0.982) | 0.992<br>(0.991 - 0.994) | 0.959<br>(0.954 - 0.964) |
| LogR | 0.847<br>(0.840 - 0.854) | 0.837<br>(0.821 - 0.853) | 0.846<br>(0.827 - 0.865) | 0.689<br>(0.677 - 0.701) |

## Notes

### Competing Interest Statement

The authors have declared no competing interest.

